# Learning to integrate? Multimorbidity, chronic care and health system adaptation in post-USAID Zimbabwe

**DOI:** 10.64898/2026.09.28.26364151

**Authors:** Fionah Mundoga, Kety Choga, Karen Webb, Patience Mangisi, Shingairai Tsvangirayi, Valient Makore, Agnes Katsidzira, Clorata Gwanzura, Tsitsi Apollo, Cleophas Chimbetete, Trudy Mhlanga, Pugie Chimberengwa, Theonevus T. Chinyanga, Efison Dhodho, Justin Dixon

**Author notes:** First joint authors (Fionah Mundoga, Kety Choga). Last joint authors (Efison Dhodho, Justin Dixon). **Correspondence to:** Fionah Mundoga, Organization for Public Health Interventions and Development, 96 Avondale West Road, Avondale. Harare, Zimbabwe.

## Abstract

Multimorbidity exposes tensions between complex health needs and health systems historically organized vertically around single diseases. While integrated chronic care has become a policy priority for many low- and middle-income countries, responses remain constrained by disease-specific configurations of knowledge, policy and practice. The USAID cuts represent a critical rupture in these arrangements, exposing dependencies while creating opportunities for integration, institutionalisation and local ownership. We examined challenges and opportunities for multimorbidity-responsive primary healthcare through a Learning Health System (LHS) lens, focusing on how information, deliberation, and action shape learning and adaptation processes in Zimbabwe during this period of transition. We conducted an ethnographic study in two urban metropolitan centers in Zimbabwe between October 2025 and February 2026, using patient journey mapping (n=23), participant observation, and in-depth interviews with healthcare workers, patients, decision-makers, health information specialists, and technical partners (n=19). We analyzed data thematically using an iterative framework informed by LHS concepts. Participants widely recognized multimorbidity as an increasingly common feature of primary care, yet care remained constrained by systems organized around individual diseases. Fragmentation was evident not only in clinical pathways but also in the information, accountability, and decision-making architectures through which health priorities were recognized and acted upon. At the same time, frontline providers routinely adapted care around patients’ multiple conditions, revealing capacity for more integrated and person-centered approaches. Existing HIV platforms, including differentiated service delivery models and regional learning networks, were frequently identified as foundations for broader chronic care integration. These findings suggest that advancing multimorbidity-responsive care requires more than service integration; it requires strengthening the learning architectures through which health priorities are defined, knowledge is generated, and action is coordinated.

## 1. Introduction

As populations age and disease patterns change globally, health systems face growing challenges in responding to multimorbidity—the co-existence of two or more chronic conditions within a single individual. Once considered a concern of high-income settings primarily, multimorbidity is increasingly recognized as a defining feature of population health across low- and middle-income countries (LMICs), where infectious diseases persist alongside rapidly rising burdens of non-communicable diseases (NCDs) (Basto-Abreu et al., 2022; Endalamaw et al., 2024; Smith et al., 2012). This challenge is particularly acute in sub-Saharan Africa, where health systems previously strengthened to address HIV, tuberculosis and malaria must now respond to syndemic interactions of chronic infectious diseases and rapidly rising hypertension, diabetes, chronic kidney disease and mental health conditions among many other rising NCDs (Dixon et al., 2025; Mendenhall et al., 2017). In Zimbabwe, the proportion of people living with HIV (PLHIV) affected by at least one NCD is projected to increase substantially between 2015 and 2035, with adult PLHIV expected to experience disproportionately high levels of multimorbidity compared to HIV-negative populations (Smit et al., 2018).

Despite growing recognition of these trends, adaptation towards integrated chronic care has been slow and uneven. A major reason for this is the enduring influence of vertical disease programmes. Critical global health scholars have long argued that such programmes, despite contributing substantially to population health gains, have reinforced disease-specific ways of organizing knowledge, resources, and care, contributing to the fragmentation and uneven resourcing of health systems (Biehl & Petryna, 2013; Prince Ruth & Marsland Rebecca, 2014). This critique extends to the metrics and accountability architectures through which global health priorities gain visibility and action (Adams 2017). Specifically, what gets counted and by whom shapes which health problems are recognized as intervention targets, often at the expense of cross-cutting categories like multimorbidity that a single indicator cannot easily measure (Dixon et al., 2023). Zimbabwe’s HIV programme is a case in point. Sustained donor investment, task-shifting and the expansion of antiretroviral therapy (ART) enabled substantial progress towards the UNAIDS 95-95-95 targets (Olpengs & Mamwa, 2025). Opportunistic infection (OI) clinics became highly effective platforms for HIV care, supported by dedicated staff, electronic information systems and reliable supply chains. Differentiated Service Delivery (DSD) models have further improved responsiveness and efficiency by tailoring care around patients’ needs through mechanisms such as fast-track medication refills, community-based ART distribution and reduced clinic attendance requirements (Ehrenkranz et al., 2020). However, the same conceptual and material infrastructures that enabled HIV programme success also helped institutionalize disease-specific ways of organizing care. A recent situation analysis (KnowM) conducted in four provinces revealed that Zimbabwe’s health system remains largely unprepared for rising multimorbidity. People living with multimorbidity (PLWMM) and those who care for them face segmented clinical spaces, long queues, inconsistent guidelines, polypharmacy, increased out-of-pocket costs, and, since NCDs (unlike HIV) remain unmanageable at the primary level, regular hospital referrals (Dixon et al., 2024). These challenges fall through gaps in parallel research, information, monitoring and evaluation (M&E) systems, and consequently remain under-recognized in policy and planning. While DSD guidance has been conceptually expanded to include other chronic conditions, implementation in practice remains at a very early stage, restricted to HIV+ clients, uneven in coverage, and hampered by continued system fragmentation (Ehrenkranz et al., 2020).

These tensions crystallizing around rising multimorbidity evidently extend beyond service integration. They expose a more fundamental and growing rift between vertical, externally driven single-disease architectures and the complex, evolving needs of whole persons, raising questions about how health priorities are defined, how knowledge is generated and mobilized, and ultimately who has the authority to shape health system responses (Abimbola, 2021). The recent disruption of USAID-supported programmes, including those for chronic diseases such as HIV and TB, has brought these questions into yet sharper relief. Among the most significant shifts in global health financing in decades, these funding changes have exposed longstanding dependencies embedded in donor-supported vertical programmes while intensifying debates around ownership, self-reliance, and health system sovereignty (Kyobutungi et al., 2025; WHO, 2025). Although often framed as a financing crisis, this moment may also be understood as a renegotiation of how health systems are governed, what is prioritized, and who decides. For countries seeking to respond to multimorbidity, the challenge is therefore not simply how to integrate services, but how to strengthen capacities for locally driven adaptation and learning amid changing health needs and shifting geopolitical realities (Sheikh et al., 2020; Sheikh & Abimbola, 2021).

Learning health systems (LHS), a conceptual framework developed at the intersection of critical global health and health policy and systems research (HPSR) by Sheikh and Abimbola, among others, has emerged as a potentially important alternative vision of health system strengthening in this regard (Dixon et al., 2026; Sheikh & Abimbola, 2021). Whereas conventional models of global health governance and health systems strengthening have often privileged standardization, upward accountability and disease-specific performance metrics, the LHS framework places greater emphasis on locally generated knowledge, iterative adaptation and learning across organizational and health system levels (Dixon et al., 2026; Sheikh & Abimbola, 2021). Multimorbidity, we have argued, provides a useful lens for examining these processes because it resists disease-specific modes of knowing and demands more holistic, adaptive, and person-centred responses (Dixon et al., 2023, 2026). In this sense, LHS can be understood not merely as a technical framework for improvement, but as an alternative approach to health system strengthening and governance—one that seeks to relocate learning, problem definition, and adaptation closer to the contexts in which health and illness are experienced and acted upon (Dixon et al., 2023, 2026). Using the conceptual lens of multimorbidity-‘learning’ health systems (Sheikh & Abimbola, 2021), this study examines how chronic care services are organized, delivered, and improved within Zimbabwean primary healthcare during a period of profound health system transition. Drawing on perspectives from patients, frontline providers, managers, policymakers, and technical partners, the study explores how disrupted funding arrangements are reshaping possibilities for integration, learning, and health system sovereignty in Zimbabwe.

## 2. Study setting and design

The study employed a participatory ethnographic design to examine how services for chronic conditions and multimorbidity are organized, delivered, and adapted in practice. Ethnographic methods captured everyday clinical routines, data practices, and decision-making processes, while ongoing stakeholder engagement informed data collection and interpretation. The research formed part of the OptiMuL programme (2025–2030), an interdisciplinary collaboration focused on establishing a learning hub to support the locally led development of integrated, adaptive services and systems for chronic conditions and multimorbidity in Zimbabwe.

### 2.1. Study setting

The study was conducted in two urban metropolitan areas, Chitungwiza (within Harare Metropolitan Province) and Bulawayo (Figure 1). Zimbabwe has a population of approximately 16.34 million in 2023 (Dixon et al., 2026), of which 38.2% live in urban areas. Urban centres have long been the epicentre of the HIV pandemic (World Health Organization, 2010) and, while notable progress has been made in HIV control, with adult prevalence (15–49 years) declining from 12.7% in 2019 to 10.5% by 2023 (Mavhunga, 2024), it continues to grapple with high rates of tuberculosis (TB) and recurrent outbreaks of waterborne diseases (World Health Organization, 2024). Simultaneously, the country now faces a significant double burden of disease, characterized by a mature HIV epidemic and a rising prevalence of non-communicable diseases (NCDs). Hypertension, diabetes, mental health conditions, and multimorbidity combinations thereof, especially HIV and hypertension, are rising rapidly, particularly in urban areas including Harare and Bulawayo, which have high rates of poverty and inequality, HIV transmission, obesity, and malnutrition. Zimbabwe’s public health system remains constrained by underfunding and systemic fragmentation. Health spending remains relatively low, with external resources making up a significant share of total expenditure (World Bank, 2022). While this has played a critical role in funding disease-specific programmes such as HIV/AIDS, TB, and malaria, the resulting fragmentation has been to the detriment of responding to NCDs and multimorbidity. It has further introduced vulnerability to funding volatility. With the cessation of US funding and related cuts in Feb-March 2025, around $522 million of aid was cancelled, accounting for about 1/3 of any year’s total health spending (Chimhete, 2025).

**Figure 1.**
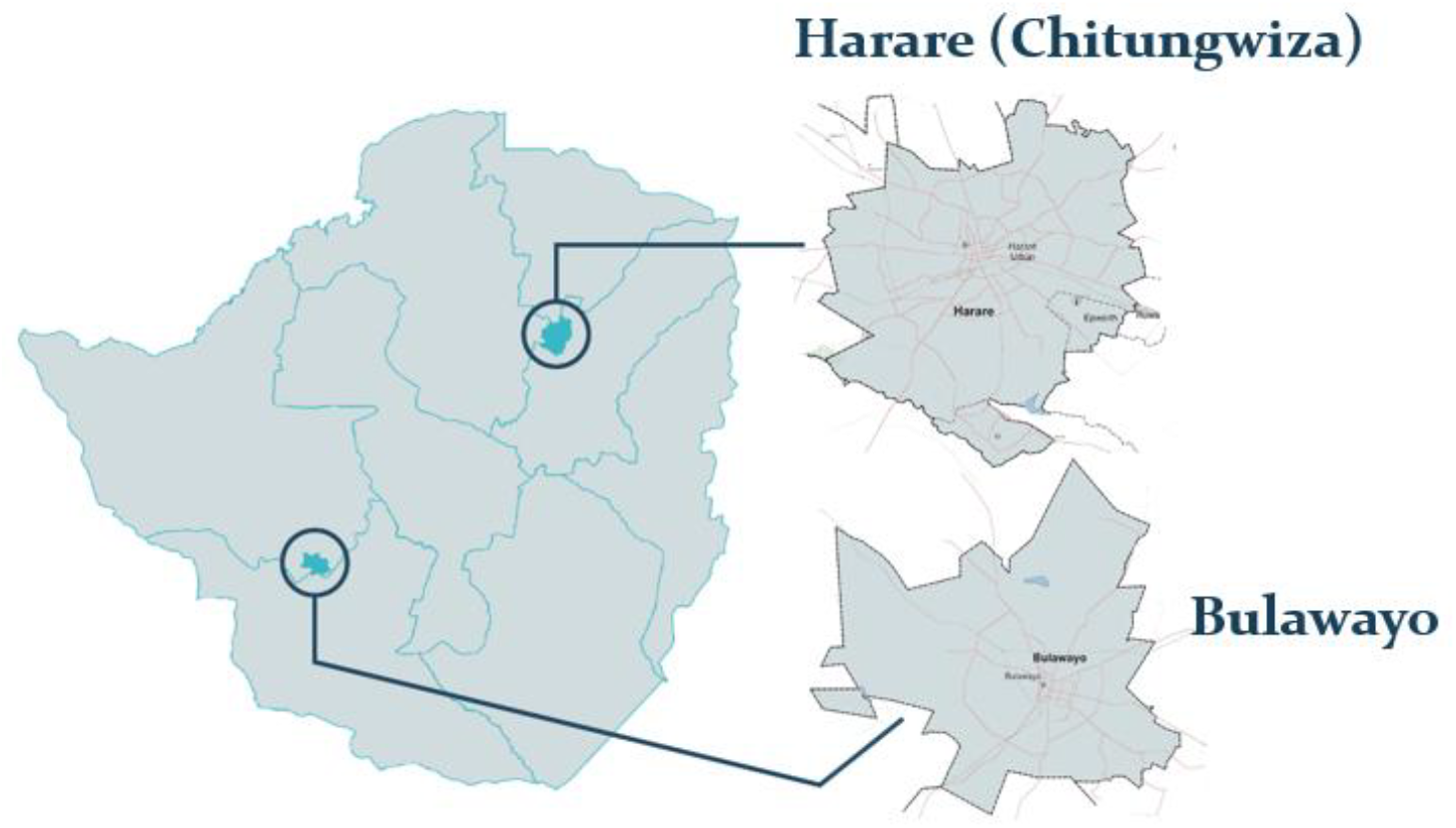
OptiMuL study settings and primary care learning sites in the metropolitan provinces of Harare* and Bulawayo. * National-level data collection occurred in Harare City, while research in primary care was conducted in the peri-urban Chitungwiza Municipality, which is within the south of Harare Metropolitan Province

Data collection took place during the first phase of the OptiMuL programme, immediately following the funding disruptions. The study was conducted in two purposively selected urban primary health clinics, Clinic 1 in Chitungwiza and Clinic 2 in Bulawayo, which served as learning sites for the OptiMuL learning hub. Both clinics were specifically selected as representative primary healthcare facilities with high patient volumes, prior experience with integrated HIV and NCD service delivery under the SANOFI Project (2022-2024) (Hove et al., 2025), and advanced implementation of the country’s Electronic Health Record (EHR), widely thought to be necessary for chronic disease service integration. Both sites are situated in high-density urban areas and have recently transitioned from intensive external support, providing a critical lens for examining the sustainability of integrated service delivery in a post-USAID funding landscape.

### 2.2. Conceptual framework

The study draws on and expands the LHS framework, a complex adaptive systems perspective that positions learning as a core function of health system strengthening (Sheikh & Abimbola, 2022). Rather than viewing improvement as the implementation of discrete interventions, LHS emphasizes continuous cycles of learning embedded in routine health system practice (Dixon et al., 2026; Jimu, 2025; Musuka et al., 2025). Within this framework, learning occurs through three means—information, deliberation, and action—and in three forms: individual, team, and (cross-) organizational whole-health system levels. To characterize the depth and type of learning, we drew on single-, double-, and triple-loop learning. Single-loop learning focuses on monitoring and improving performance against existing goals (e.g., M&E systems); double-loop learning involves questioning and adapting underlying assumptions, priorities, and ways of working; while triple-loop learning, or ‘learning to learn’, concerns strengthening the conditions and capacities through which learning itself occurs (Sheikh & Abimbola, 2021). In this study, we used LHS as an analytic lens to examine how information, deliberation, and action are configured to enable—or constrain—health system responses to multimorbidity in Zimbabwe. Building on previous work that adapted the LHS framework to multimorbidity and integrated chronic care (Dixon et al., 2026), we explored the extent to which existing learning processes remained organized around individual diseases versus supporting more person-centred and adaptive responses to complexity (**Figure 2**). Drawing on findings from the KnowM situation analysis and broader critical global health and LHS scholarship, we hypothesized that **Figure 2a** reflects Zimbabwe’s prevailing chronic care configuration, characterized by fragmented, disease-specific, and predominantly single-loop learning architectures. In contrast, **Figure 2b** illustrates a potential future in which disrupting established funding arrangements creates opportunities for more integrated learning, governance, and care.

**Figure 2.**
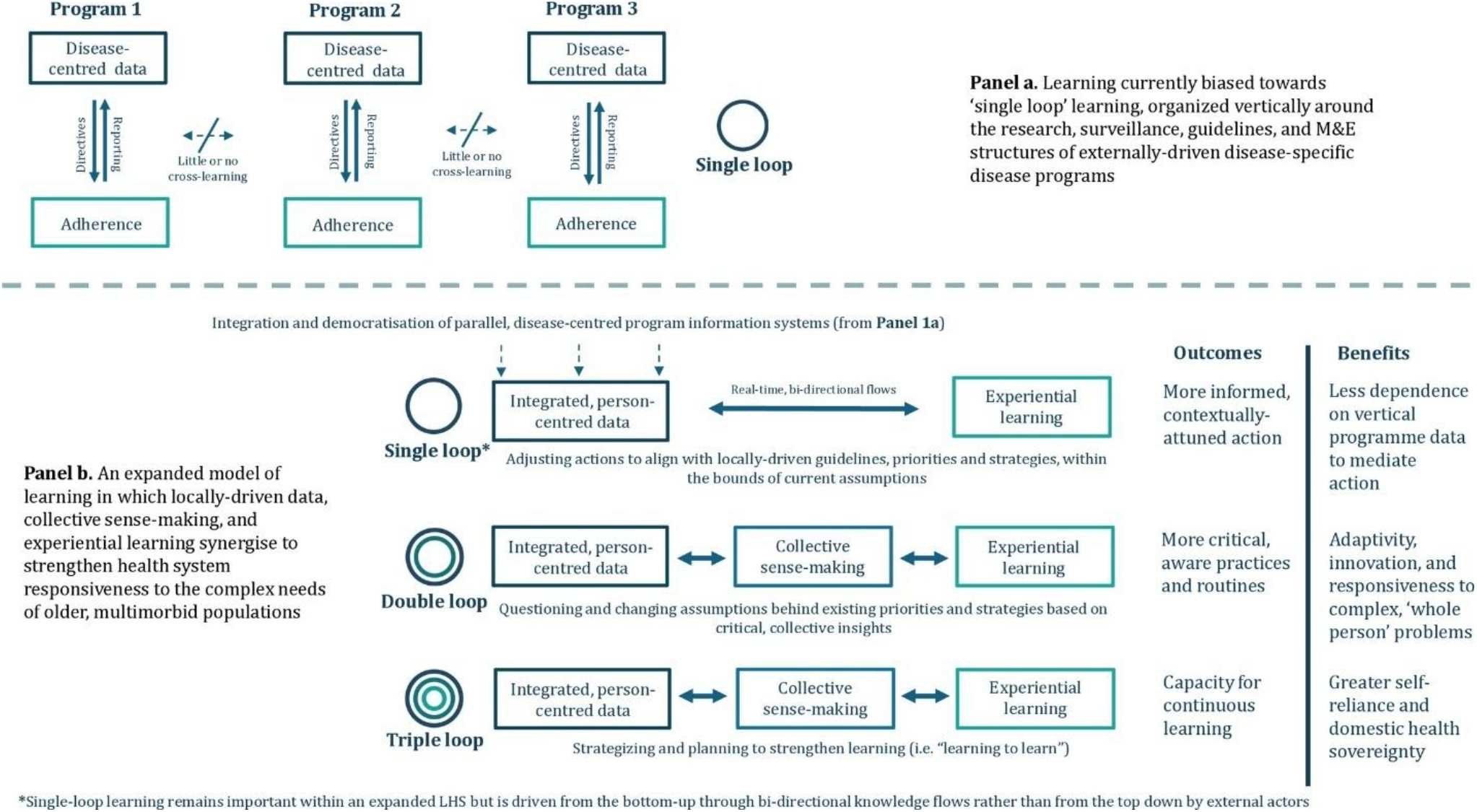
Conceptualizing the means, loops and benefits of learning relative to current health system architectures built around vertical disease programmes (Dixon et al., 2026).

### 2.3. Data collection

Data collection took place between October 2025 and February 2026. Within a participatory ethnographic design, data collection methods included observations, patient journey mapping, and stakeholder interviews.

#### 2.3.1. Patient journey and care pathway mapping

Starting from the bottom up, we first conducted patient journey mapping – a qualitative technique used to visualize and analyse the experiences of individuals as they navigate a complex system (Bulto et al., 2024). We chose this method to provide a rich, patient-centred understanding of clinical workflows, data flows, and structural barriers affecting multimorbidity care in practice. It was based on a modified Service and Readiness Assessment (SARA) survey conducted during the prior KnowM study, which gave us a basic understanding of clinical space, staffing, medicines, equipment, service delivery points, guidelines, information systems, referral pathways, and private sector partnerships (Dixon et al., 2024). We updated these findings before commencing patient journey mapping. Data from journey mapping were collected as field notes through direct, non-participant observation of patients known to have hypertension, HIV, or both multimorbid HIV-hypertension; (n=23). FM and KC shadowed individual patients from arrival to departure, meticulously documenting each step of their journey. All patients provided written informed consent before taking part. Observations were captured in real-time using a structured template (provided as Supplementary File 1) that included:

- Timestamps – to measure wait times and the duration of each interaction.
- Locations – to map the physical movement of the patient across different service points (e.g., reception, triage, consultation room, pharmacy).
- Provider Interactions – to document the nature of clinical and administrative encounters.
- Data and Information Flow – to track how patient data was recorded, retrieved, and utilized across different paper-based and electronic systems at the point of care (e.g., patient booklets, registers, EHR).

#### 2.3.2 Observation of learning Processes and arenas

Widening the scope beyond clinical workflows, we used targeted participant-observation to ‘follow’ patient data and other information sources across the data-deliberation-action pathway: from facility data retrieval, consolidation, and use; to upward reporting, analysis, and feedback mechanisms; to (sub)national review, M&E, technical and decision-making meetings; and finally capturing the various ways in which decisions, policies, interventions, or adaptations were put into practice and iterated on the ground. ED and JD, supported by FM and KC, spent focused periods of time (spanning from one hour to 3-5 days) with purposively selected individuals with specialist knowledge of data systems and health systems organizational structures, shadowing their everyday routines and doing walk-throughs of software, databases, and flow diagrams/visual representations. We also sat in on – and, where appropriate, took part in various meetings, trainings, and reviews; followed up where necessary with stakeholder interviews (detailed in 2.3.3). This ‘wide-angle’ approach enabled us to identify how information was generated, shared, and used across different levels of the health system, as well as the opportunities, constraints, and dependencies shaping learning, adaptation, and decision-making for multimorbidity care.

#### 2.3.3. In-depth stakeholder interviews

Dovetailing with the observational work, we conducted in-depth interviews with key stakeholders across the health system, including expert patients, health workers, informaticians, decision-makers, technical partners, and academic researchers (many of whom were also part of other research activities). In total, we conducted 19 interviews. FM, KC, ED, and JD conducted these interviews using role-specific topic guides tailored to each individual, and shared high-level themes and insights from previous activities. Written informed consent was obtained from all participants before the interview; interviews lasted between 60 and 90 minutes and were audio-recorded where permitted; if not, field notes were taken instead. Audio-recordings were transcribed for analysis.

**Table 1.** Participant demographics and activity breakdown.

| **Participant category** | **In-depth interviews** | **Participant observation** | **Patient journeys** | **Total** |
| --- | --- | --- | --- | --- |
| **People living with multimorbidity** | - | 11 | 11 | **11** |
| **People living with HIV only** | - | 6 | 6 | **6** |
| **People living with hypertension only** | - | 6 | 6 | **6** |
| **Health professionals (Clinics 1 and 2)** | 7 | 4 | - | **11** |
| **Decision makers within national MoHCC** | 5 | - | - | **5** |
| **Health informatics experts** | 2 | - | - | **2** |
| **Technical partners/NGOs** | 3 | - | - | **3** |
| **City Health Department (Bulawayo and Chitungwiza)** | 2 | 1 | - | **3** |
| **Total** | **19** | **28** | **23** | **46** |

\*Participants enrolled for patient journeys were the same ones that were observed during care.

### 2.4 Data analysis

All interview transcripts and observation notes were uploaded into NVivo 15 for data management and analysis. We used thematic analysis, coding both deductive codes derived from the LHS framework and inductive codes generated from the data. JD and FM analysed data as an ongoing, iterative process and fed it back into ongoing data collection. Preliminary findings were shared at various fora with the learning hub and the Ministry of Health and Child Care (MoHCC) departments to aid and refine interpretations. The team triangulated all data sources to develop final findings, interpretations, and recommendations.

## 3. Findings

Drawing on in-depth interviews, observations, and patient journey mapping, the results are organized around four main themes: service delivery context; the ‘wake-up call’ of the United States (US) cuts; centring experiential learning in service integration; and the longer-term need to strengthen the learning-enabling environment for sustainable, adaptive multimorbidity care.

### 3.1 Service delivery context

#### 3.1.1 Fragmented chronic and multimorbidity services

Across both clinics, healthcare workers routinely managed multiple chronic conditions within a single encounter, particularly in outpatient (OPD) and OI services. Multimorbidity was widely described as becoming the norm rather than the exception in primary care, with combinations of HIV, hypertension, diabetes and mental health conditions especially common. Participants linked this trend to improved HIV survival and population aging, noting that the primary health risks faced by many people living with HIV are now increasingly driven by unmanaged NCDs:

*“*…*We are now going to have all conditions in one patient. You will have a 70-year-old woman or man who has diabetes, is overweight, has hypertension, arthritis, and is starting to suffer from cancer*…” [Decisionmaker_07_Byo]

*‘People with HIV are the ones with most NCDs…so they’re dying more of NCDs than HIV*…’[HCW_03_Byo]

Despite its growing prevalence, multimorbidity remained poorly accommodated within service delivery and information systems organized around individual diseases. Observations revealed parallel registers, EHR modules and patient pathways that often required individuals to navigate multiple queues, consultations and pharmacy systems for related conditions within a single visit. Participants described these arrangements as a legacy of vertical programme structures that continue to shape guidelines, reporting systems and resource allocation:

*‘The other problem is from a patient perspective…this whole siloed approach is bad because then they have a whole bunch of pills* [Decisionmaker_04_Harare]

#### 3.1.2. Early integration efforts and its challenges

Both clinics had been the focus of early efforts to integrate chronic care. As EHR-optimised sites, they had implemented digital systems intended to support longitudinal care, while national and partner-led initiatives had introduced integrated screening and service delivery models. For example, the HIV programme increasingly incorporated screening for hypertension, diabetes and mental health conditions, and its Operational and Service Delivery Manual (OSDM) had begun to formalize these approaches through DSD models and routine NCD screening for people receiving HIV care. Nurses described routinely screening for NCDs during HIV visits, providing counselling across conditions, administering medication and making referral decisions based on clinical judgement. At Clinic 2, for example, OPD nurses commonly managed patients with combinations of HIV, hypertension, diabetes and asthma within a single consultation, reflecting considerable frontline capacity to adapt to emerging multimorbidity realities. As one nurse explained:

*“Say today they are coming for their hypertension medication… coming for their ART supply… want… diabetic refill…”* [HCW_04_Byo]

Despite these developments, integration efforts were widely viewed as being at an early stage and largely confined to HIV-positive populations, with limited formal integration between OI and OPD services. Participants noted that the EHR system continued to mirror disease-specific funding and reporting structures through separate HIV, TB, and chronic disease modules, reinforcing rather than overcoming programme silos. Consequently, much of the day-to-day integration work depended on facility-level initiative and responsiveness to patient needs rather than formal policy or system redesign. Healthcare workers described blending services as a practical necessity, while decision-makers highlighted how professional identities shaped through years of vertical programming continued to constrain more holistic approaches to care. As one decision-maker observed:

*‘But because their mind says I am an OI Nurse, it means this is somebody else’s business. Go see the doctor and then the doctor will give you the management that you need. Even though I can make this diagnosis, give you the proper prescription that’s going to control your hypertension, I am an OI Nurse’* [Decisionmaker_06_ Harare]

Successful integration was therefore seen to require not only changes to service delivery models, but also a shift from disease-specific roles towards more generalist approaches to chronic care. Overall, multimorbidity care was gradually becoming integrated into everyday practice, but remained heavily dependent on local adaptation and constrained by the enduring legacies of vertical programming.

### 3.2 Wake-up call: rapid institutionalisation and integration

Decision-makers described the 2025 cessation of global health funding as an unprecedented shock that extended beyond financing to governance, workforce stability and service continuity. Programmes that had historically been among the most well-resourced suddenly faced severe financial pressure, resulting in staffing disruptions, programme stoppages and increased demand for cross-programme coordination:

*“What has happened in 2025 was really unprecedented… It created some shocks to the system…that increased demand for cross-programme learning and coordination”* [Decisionmaker_11_Har]

At City and facility level, the stop-work orders translated into immediate human resource and commodity shortages, with interruptions in medicine supply generating anxiety among both healthcare workers and patients. One healthcare worker described how reductions in (Antiretroviral Therapy) ART dispensing intervals prompted concerns about future treatment availability:

*‘We used to give six months, then we started to give three months. And the community, they start to react now. They say in a panic mode, and they think that the medication is no longer there’ [HCW_06_CHIT]*

Participants argued that the abrupt transition exposed longstanding dependencies within donor-supported programmes and highlighted the fragility of reforms that remained incomplete. Several described the disruption as occurring at a critical moment in ongoing efforts to strengthen digital systems, workforce capacity and service integration:

*‘So, the problem was the abrupt disruption…right at the point where we thought we had significant system maturity to start a significant transition*…*you are rolling a big rock uphill…if you lose the momentum it will roll back on you’* [Decisionmaker_04_Har]

At the same time, the funding transition was widely framed as a catalyst for institutionalisation and greater domestic ownership. Technical partners described accelerating efforts to transfer competencies and responsibilities into the public health system, while MoHCC stakeholders viewed the disruption as a wake-up call that clarified the need for stronger local ownership of core health system functions:

*‘With the cuts…big change, needed us to think in other terms so had to accelerate the health systems strengthening agenda, rebrand that to institutionalisation and quickly transfer competencies to the health system’* [TechPartner_01_Har]

*‘The [EHR] system is now too big to fail… what has happened now accelerates the reason. The need for us to just own the space. Not crowding out contributors, but we need to own the core implementation of this thing’* [Decisionmaker_04_Har]

Beyond institutionalisation, participants also saw opportunities to advance integration by leveraging infrastructure developed through the HIV response, including financing mechanisms, DSD models and regional learning platforms such as the HIV Coverage, Quality, and Impact Network (CQUIN). Several argued that existing HIV platforms could provide a foundation for broader chronic care approaches that extend beyond HIV and support people living with multimorbidity. Proposed reforms included expanding financing arrangements and governance structures to support integrated chronic care rather than disease-specific programmes:

*‘…it already has structures. I think it would be a nice, an easy way, like a low-hanging fruit, to say you put all the money in NAC [National AIDS Council] and then expand the scope and mandate of NAC’* [Decisionmaker_07_Byo]

Overall, the post-USAID transition was experienced as both disruption and reorientation. While destabilizing in its immediate effects on staffing, medicines and service delivery, it also sharpened calls for institutionalisation, greater domestic ownership and more integrated approaches to chronic care organized around people rather than individual diseases.

### 3.3 The longue durée: building a sustainable learning-enabling environment for integrated, adaptive, person-centred care

While participants frequently discussed immediate funding, workforce and policy challenges, they also reflected on deeper system capacities and vulnerabilities shaping the long-term response to multimorbidity. These centred on how information is generated and used, how actors collectively make sense of emerging challenges, and how experiential learning can be translated into service improvement.

#### 3.3.1 Integrating and democratising health information

Participants widely viewed EHR as a major opportunity for strengthening chronic and multimorbidity care. Designed to replace fragmented paper registers and disease-specific reporting systems, EHR promised a unique patient identifier, reduced duplication and a longitudinal view of patient care that could support coordinated decision-making across conditions. At both clinics, EHR was already embedded in registration, consultations, pharmacy management and HIV care, while a chronic care module had recently been introduced for long-term NCD management. However, participants consistently described the system as reproducing vertical-programme logic rather than overcoming it.

Although HIV and chronic disease modules existed within the same platform, they remained largely disconnected. Clinicians reported that multimorbidity was often visible only through individual recall, patient disclosure, or manual cross-checking between modules:

*‘For the electronic health records, there’s no integration. The chronic disease, they have their own module, the HIV, they have their own module, so it’s up to the clinicians to know that this patient also has [an NCD]*. [HCW_03_Byo]

Decision-makers similarly argued that current EHR structures mirrored disease-specific funding and reporting systems, limiting the visibility of multimorbidity and constraining coordinated care:

*‘Our systems are also supposed to be integrated, and I think it’s one of our current major limitations with our electronic health record system that everything is still in modular form. You know, the HIV module, the TB module, this module, this module. The best scenario would be, you know, having all the information at once*’ [Decisionmaker_06_ Harare].

Participants further noted that existing systems generated few forward-looking outputs, such as synchronized appointments, patient line lists, or integrated population views. Consequently, multimorbidity remained clinically common but analytically obscured. Yet the post-USAID transition was also seen as creating momentum to redesign digital infrastructure around patients rather than programmes. Across stakeholder groups, the preferred future state was a longitudinal chronic care record in which HIV became one component of a broader patient profile rather than a standalone category:

*‘The first thing a system must do is follow the patient and not the disease*’ [Decisionmaker_04_ Harare].

*‘If I am to click on [a patient], what should come up is all my demographics, whether it was management for hypertension or for HIV. I should be able to go through all those conditions, take hypertension, see management, how are you doing and your current BPs, what actions do I need to take, move to the next, or you also have this. And then I manage those. Instead of just looking at a name, getting the demographics, and then going, we are in OI, so I’m going to click the OI module, and there’s nothing else that’s there’* [Decisionmaker_06_Harare]

In this framing, EHR was viewed not simply as a record-keeping tool, but as a potential backbone for integrated chronic care, capable of improving visibility of multimorbidity, supporting differentiated service delivery, and enabling more coordinated, person-centred care.

#### 3.3.2. Collectively ‘making sense’ for coordinated action

Alongside information systems, participants emphasized the importance of deliberative spaces through which data could be interpreted and translated into action. At facility level, routine review meetings were described as important mechanisms for identifying bottlenecks, monitoring performance and coordinating service improvements. However, these deliberative processes largely reflected the same programme-specific architecture that characterized information systems. HIV indicators were routinely generated, reviewed, and acted upon, while NCD and multimorbidity data were often incomplete, manually aggregated, or absent altogether:

*‘The chronic module does not appear. Because they just register those patients, just register the name and then the prescription. So, when then we have the reports, those patients do not appear’* [HCW_01_CHIT]

Participants linked these gaps to parallel paper and EHR systems and the underdevelopment of the chronic module’s reporting capabilities, limiting the ability to generate integrated views of multimorbidity care. While HIV indicators were automatically generated and regularly reviewed, with programme representatives continuously visiting, requesting, and checking the quality of specific indicators of interest, NCD indicators required manual aggregation, were rarely chased, or were not systematically reported. As a result, multimorbidity, although clinically common, remained analytically obscured.

As information moved upwards through district, city and national structures, participants described increasingly fragmented learning arrangements organized around disease programmes, directorates and funding streams:

“*What then came through over the years was verticalization… taking one area of knowledge, one disease condition, then you say, strictly stick to this because that’s what is funded” [Decisionmaker_02_Har]*.

At the same time, the US cuts were seen to offer new possibilities for creating new deliberative platforms across conditions, programmes, and sectors, in a way that vertical programmes had structurally inhibited:

*Policy, strategy, and operational guidelines almost fall under the same banner of policy …they have a lot of intersections and overlap. They can be worked around together rather than as separate areas* [Decisionmaker_06_Har]

The most notable example was the emergence of a CQUIN integration maturity dashboard, adapted from existing DSD learning platforms to support a broader chronic care approach irrespective of HIV status (CQUIN, 2025). Participants viewed this as an important step toward more integrated learning and accountability. Several also proposed dedicated learning platforms or “think tanks” that could bring together actors across programmes, sectors and levels of the health system to support the ongoing refinement of integrated chronic care.

### 3.4 Centring experiential learning in service integration

Participants consistently framed integration not as a discrete intervention but as an ongoing process of adaptation across actors, resources and contexts. While national policies could define broad integration goals and service delivery options, respondents argued that those directly involved in care delivery should shape implementation decisions. The challenge was therefore not simply determining what integrated care should look like but creating mechanisms through which frontline experience could inform how integration is designed, tested and refined over time.

Facilities that had already achieved some degree of integration—through research partnerships, technical assistance or local initiative—were widely viewed as valuable learning sites. Participants argued that these settings could help identify which organizational arrangements were most appropriate under different circumstances and support iterative adaptation as conditions changed. Quality improvement (QI), particularly the MoHCC’s Kaizen/5S approach, was frequently identified as an underutilized mechanism for this process. Unlike externally funded pilot projects, QI was described as embedded within routine practice and therefore well suited to addressing the everyday challenges of multimorbidity care:

*‘Somebody who knows what they do every day is in a better position to improve their work*…*than somebody from the head offices who comes to the clinic’* [Decisionmaker_03_Harare]

Participants viewed QI as a practical means of identifying bottlenecks, testing locally feasible solutions and continuously refining integrated care within existing constraints. In this sense, QI was positioned not merely as a technical tool, but as an emergent governance mechanism through which integration-as-process could become embedded within everyday health system practice rather than remaining dependent on externally funded initiatives.

## 4. Discussion

Using the conceptual lens of multimorbidity-learning health systems (Figure 2), this study examined how chronic care services are organized, delivered and improved within Zimbabwe’s primary healthcare system during a period of profound health system transition. Our findings suggest that multimorbidity is not merely a deficit of information but a consequence of how health systems are organized to produce, interpret and act on information. In this sense, multimorbidity functions as what Biehl & Petryna (2013) describe as a challenge to established regimes of visibility and intervention, exposing the limits of disease-specific ways of knowing and governing health (Adams, 2017; Biehl & Petryna, 2013; Dixon et al., 2023; Reubi, 2018). Participants repeatedly described information, reporting, and decision-making systems that made multimorbidity difficult to recognize, measure, and act on despite its growing prominence in everyday clinical practice. Patient journey mapping further demonstrated that fragmentation was experienced not simply as an organizational problem but as an everyday burden involving multiple queues, consultations, appointments, and medication pathways. Patients frequently moved between service points, providers and record systems in ways that increased treatment burden and the risk of missed care. These findings reinforce wider concerns that disease-specific models of care often transfer the work of integration from health systems to patients and their families (Ameh et al., 2017; Dixon et al., 2023). However, viewed through a multimorbidity-LHS lens, the findings also point beyond critique. Across interviews, observations and patient journeys, we observed important forms of adaptation through which healthcare workers, managers and decision-makers sought to bridge these divides, revealing latent capacities for integration already present within the system. Rather than representing only a challenge to existing arrangements, multimorbidity may therefore serve as a focal point around which more integrated, adaptive and locally driven forms of learning and care can emerge.

Consistent with our conceptual framework, the findings suggest that chronic care in Zimbabwe remains organized around the fragmented, disease-specific and predominantly single-loop learning architecture represented in **Figure 2a**. While participants recognized the need for more integrated and person-centred approaches, information systems, monitoring structures and accountability mechanisms remained primarily oriented towards measuring performance against existing programme-specific targets rather than generating actionable knowledge about multimorbidity or chronic care more broadly. In other words, learning was largely focused on improving performance within established disease-specific architectures rather than questioning whether those architectures remained fit for purpose. Although EHR has expanded substantially in Zimbabwe and are widely viewed as important enablers of integration (Mugauri & Chimsimbe, 2025), participants described systems that continue to mirror the priorities and reporting requirements of historically vertical HIV, TB and disease-specific programmes. As a result, multimorbidity—and complexity more generally (Dixon et al., 2025; Sturmberg et al., 2021)—remains poorly visible within routine information systems despite becoming increasingly common in everyday clinical practice. This invisibility matters because it constrains health systems’ ability to recognize, learn from, and respond to emerging patterns of need and complexity.

At the same time, the findings identified important examples of the more adaptive, multimorbidity-responsive learning architecture shown in **Figure 2b**. These were most evident in double-loop learning, where actors questioned and adapted underlying assumptions in response to changing realities and needs (Sheikh & Abimbola, 2022). Healthcare workers routinely described multimorbidity as the norm rather than the exception and often adapted service delivery accordingly, crossing formal programme boundaries to manage multiple conditions in a single consultation. Such practices challenge longstanding distinctions between HIV, NCD and other chronic disease services and suggest that many actors are already beginning to reframe chronic care around patients rather than diseases (Ameh et al., 2017). Importantly, this questioning was not confined to frontline providers. Participants across the health system—including managers, policymakers and technical partners—described growing recognition that existing disease-specific arrangements were increasingly misaligned with the realities of multimorbidity and chronic care. Similar processes have been conceptualized as forms of collective sensemaking through which actors reinterpret problems, challenge established assumptions and develop new ways of responding to changing conditions (Gilson et al., 2021). Examined through the LHS framework, these findings indicate that although disease-specific learning architectures continue to predominate, practices of reflection, adaptation, and reframing are already developing across multiple levels of the health system and could serve as the basis for more integrated and responsive models of chronic care.

The abrupt cessation of USAID-supported programmes created a particularly important context in which these assumptions and dependencies became visible – or, at least, could no longer be avoided (Kyobutungi et al., 2025). Rather than simply exposing funding shortages, the disruption made visible relationships and dependencies that had become normalized over decades of donor-supported health system development. Participants repeatedly described the funding disruptions as exposing vulnerabilities that had accumulated over decades of reliance on externally funded vertical programmes, with immediate consequences including workforce disruptions, commodity shortages and interruptions to planned reforms. However, participants also framed the transition as a moment to reflect on ownership, institutionalisation, and long-term sustainability. Instead of interpreting the post-USAID context merely as a crisis or an opportunity, our findings suggest it is better understood as a critical rupture: a moment that simultaneously destabilized existing arrangements and prompted collective questioning of how chronic care should be organized, governed and improved (Kyobutungi et al., 2025; Ntusi, 2025). In this sense, the significance of the funding transition extends beyond resource mobilization alone. It has brought renewed attention to questions of health system sovereignty, including who defines priorities, what forms of knowledge are valued, and where responsibility for adaptation and improvement resides (Ntusi, 2025). Similar debates are increasingly emerging across Africa as countries seek to navigate shrinking external funding envelopes while responding to growing burdens of chronic disease and multimorbidity (Kyobutungi et al., 2025). From an LHS perspective (Dixon et al., 2026; Sheikh et al., 2022), these developments represent not simply a funding shock, but a system-wide double-loop learning moment in which previously taken-for-granted assumptions about vertical programming, donor dependence, and disease-specific governance are being actively reconsidered.

Viewed through the lens of triple-loop learning, the central question becomes how to sustain, strengthen, and institutionalize the emerging learning architecture shown in **Figure 2b** across the health system. Whereas double-loop learning involves questioning existing assumptions and arrangements, triple-loop learning focuses on developing enduring capacities to learn, adapt, and improve amid ongoing uncertainty and complexity (Sheikh et al., 2022). If the USAID transition has exposed the limitations of existing chronic care architectures and stimulated new ways of thinking about integration, ownership and service delivery, attention must now turn to creating the conditions through which such adaptation can be sustained beyond the current moment of disruption. In this sense, triple-loop learning shifts attention from reforming existing arrangements towards strengthening the infrastructures, relationships and capabilities through which learning itself occurs (Sheikh et al., 2022). Our findings point to several priorities, including integrated and patient-centred information systems, deliberative platforms that connect actors across programmes, professions and sectors, and practical learning environments where new models of chronic care can be tested, adapted and refined. Participants emphasized collaborative learning platforms that move beyond fragmented technical working groups toward more action-oriented engagement. Existing HIV-related networks, particularly the CQUIN network (CQUIN, 2025), were frequently cited as capacities worth leveraging and expanding beyond HIV, alongside adaptive service delivery approaches such as DSD. Importantly, participants suggested that this transition is already underway. The CQUIN network and its integration maturity framework have recently been adapted to support broader chronic care integration efforts, including the development of integration staging and dashboard approaches that extend beyond HIV services (CQUIN, 2025). Participants valued these platforms not simply for sharing information but for supporting collective interpretation of emerging challenges, peer-to-peer learning and the iterative refinement of integrated chronic care. Such mechanisms may be particularly important in the post-funding transition context, where more immersive and less hierarchical forms of engagement between those who generate knowledge and those responsible for implementation may become necessary rather than optional. These approaches to learning are interdependent rather than distinct capabilities; information must feed deliberation, deliberation must shape action, and the consequences of action must produce new information to support cycles of learning and adaptation (Sheikh et al., 2022). Viewed through a multimorbidity-LHS lens, these developments suggest that the foundations of a more integrated and adaptive learning architecture already exist; the challenge is to strengthen and connect them in ways that support continuous learning across conditions, programmes, and levels of the health system.

Organized around the means and loops of learning presented in **Figure 2**, the key priorities and implications emerging from this study are presented in **Box 1**.

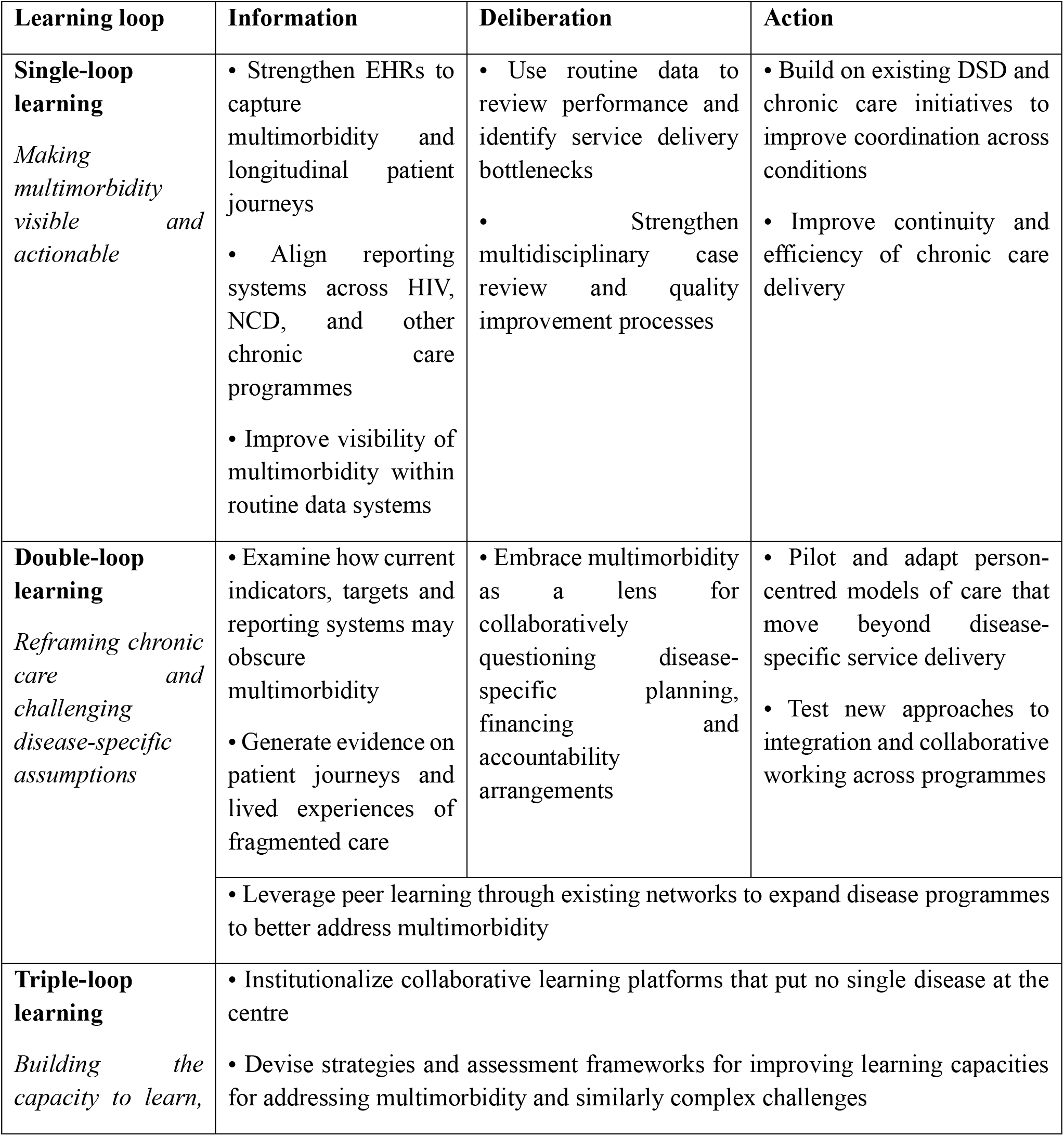
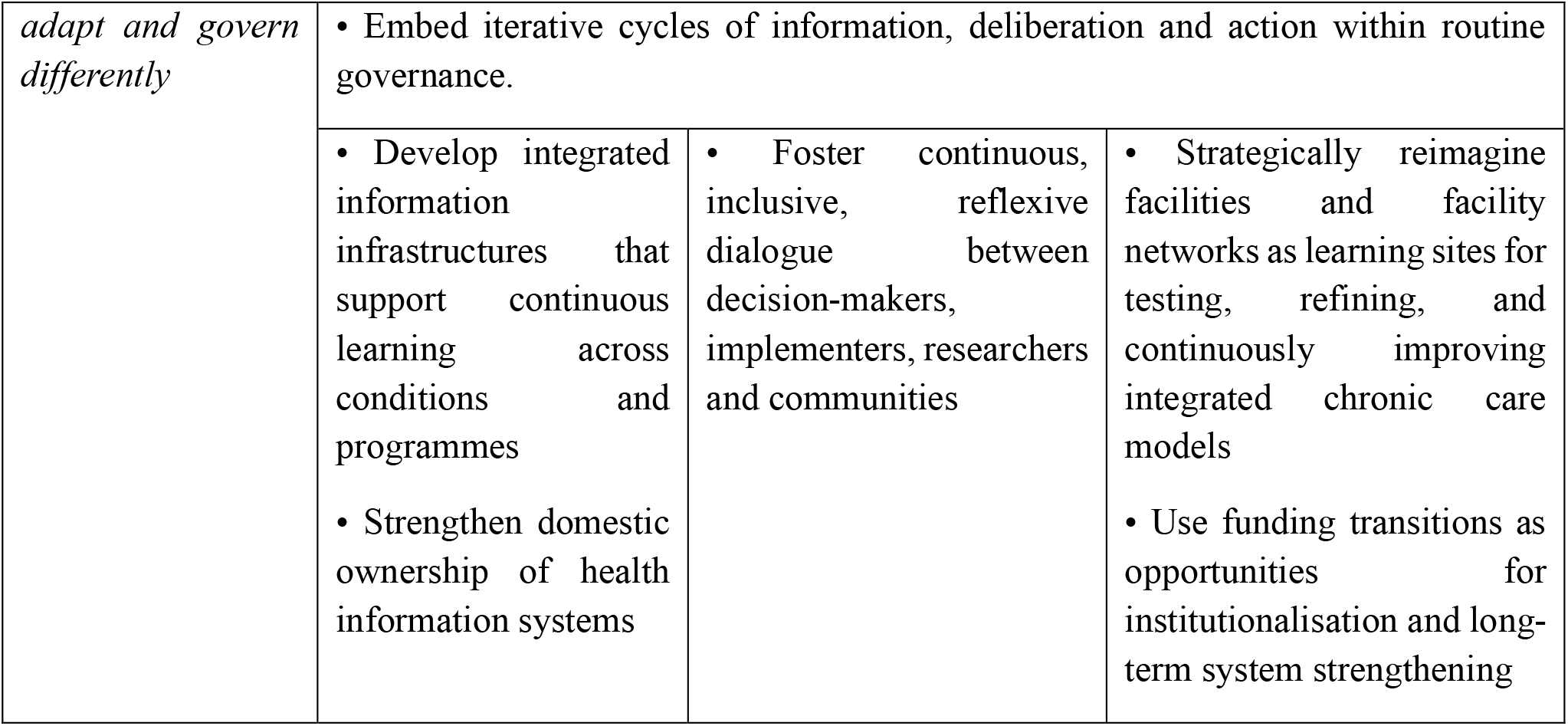

## 4. Limitations and strengths

Our study has several limitations. First, we collected data in two urban primary healthcare clinics with relatively advanced EHR implementation and prior exposure to integration initiatives, which may limit transferability to rural or less-resourced settings. Second, the rapidly evolving funding environment means that some organizational responses to the USAID cuts remain emergent and incomplete. Nevertheless, the timing of the study immediately following the funding disruptions provided a rare opportunity to examine how frontline actors, managers, and decision-makers interpret and respond to abrupt systemic change in real time. Finally, while this study focused primarily on organizational and health systems dimensions of multimorbidity care, future work should further explore the social, economic, and household impacts of fragmented chronic care pathways among people living with multimorbidity.

## 5. Conclusion

Viewed through a learning health systems lens, multimorbidity highlights not only the need to integrate services, but also the need to reconfigure how health systems generate knowledge, deliberate on priorities and coordinate action. The post-USAID transition has exposed the limitations of externally driven disease-specific architectures while creating opportunities for more integrated, adaptive and locally governed systems of chronic care. Whether these opportunities are realized will depend on the extent to which countries can strengthen the infrastructures, relationships, and learning capacities required to respond to complexity in ways that are person-centred, contextually grounded, and increasingly self-reliant. Beyond redesigning services, integration requires an institutional learning architecture that connects information, deliberation, and action.

## Supporting information

Supplemental File 1_Patient Journey Mapping

## Data Availability

All data produced in the present study are available upon reasonable request to the authors

## CRediT authorship contribution statement

**Fionah Mundoga**: Methodology, data curation, formal analysis, investigation, methodology, writing-original draft, writing-review and editing. **Kety Choga**: Methodology, project administration, writing-review and editing. **Karen Webb:** Funding acquisition, project administration, resources, writing-review & editing. **Patience Mangisi**: Resources, writing-review and editing. **Shingairayi Tsvangirai:** Resources, writing-review and editing. **Valient Makore**: Resources, writing-review and editing. **Agnes Katsidzira**: Investigation, Writing-review and editing. **Clorata Gwanzura:** Writing-review and editing. **Tsitsi Apollo**: Writing-review and editing. **Cleophas Chimbetete:** Writing-review and editing. **Trudy Mhlanga**: Writing-review and editing. **Pugie Chimberengwa:** Writing-review and editing. **Theonevus T. Chinyanga**: Funding Acquisition, Project administration; Resources, Writing-review and editing. **Efison Dhodho:** Conceptualization, formal analysis, data curation, formal analysis, investigation, methodology, writing-review and editing, resources, supervision. **Justin Dixon:** Conceptualization, formal analysis, data curation, formal analysis, investigation, methodology, writing-review and editing, resources, supervision,

## Ethics Statement

The study was approved by the Medical Research Council of Zimbabwe (ref. MRCZ/A/3315), the London School of Hygiene & Tropical Medicine (ref. 31515) and the local authorities responsible for the participating primary care clinics.

## Data Availability Statement

Data will be made available upon request.

## Acknowledgements

We gratefully acknowledge the patients, healthcare workers, policymakers, and other stakeholders who generously shared their time and experiences with this study. We also thank the participating health facilities for their support in facilitating the research and the wider OptiMuL research team for their contributions and support throughout the study.

