## Supplemental File 1_Patient Journey Mapping for "Learning to integrate? Multimorbidity, chronic care and health system adaptation in post-USAID Zimbabwe"

**Appendix 1**

**Supplementary File 1: Participant Journey Mapping Template**

PATIENT PROFILE

| | |
|---|---|
| **Observer Name:** | |
| **Date of Observation:** | ____/____/________ |
| **Patient Study ID:** | NKETA-2025-_______ |
| **Patient Age:** | _______ years |
| **Patient Sex:** | ☐ Male ☐ Female |
| **Known Conditions:** | ☐ HIV ☐ HTN ☐ Diabetes ☐ TB ☐ Other: __________ |
| **Visit Type:** | ☐ Routine ☐ Unscheduled ☐ New diagnosis ☐ Follow-up |
| **Condition Cluster:** | ☐ HIV in care<br>☐ HTN in care<br>☐ HIV newly diagnosed with HTN<br>☐ HTN/HIV in care |

1. SWIMLANE 1: PATIENT JOURNEY

| Step # | Time Start | Time End | Patient Action/Experience | Location | Burdens/Notes |
|---|---|---|---|---|---|
| **1** | | | Arrives at clinic | Gate/Entrance | |
| **2** | | | Joins queue for triage | Triage | Wait time: ___ min |
| **3** | | | Vital signs taken (BP, weight, temp) | Triage | |
| **4** | | | Moves to OPD/ART/NCD clinic | | |

| | | | | | |
|---|---|---|---|---|---|
| **5** | | | Joins queue | OPD/ART/NCD | Wait time: ___ min |
| **6** | | | Consults with clinician | Consultation Room | |
| **7** | | | Sent for lab tests | Laboratory | |
| **8** | | | Joins lab queue | Laboratory | Wait time: ___ min |
| **9** | | | Bloods drawn/sample given | Laboratory | |
| **10** | | | Returns to clinician with results | Consultation Room | |
| **11** | | | Receives prescription | Consultation Room | |
| **12** | | | Goes to pharmacy | Pharmacy | |
| **13** | | | Joins pharmacy queue | Pharmacy | Wait time: ___ min |
| **14** | | | Receives medication | Pharmacy | |
| **15** | | | Receives next appointment date | Appointment Desk | |
| **16** | | | Exits clinic | Gate/Exit | Total time: ___ hrs |

2. SWIMLANE 2: CLINICAL SERVICE PROVIDER WORKFLOW

| **Step #** | **Time** | **Provider Action** | **Provider Cadre** | **Service Point** | **Decision? (Y/N) & Notes** |
|---|---|---|---|---|---|
| **1** | | Greets patient, takes vital signs | Nurse/HCA | Triage | |
| **2** | | Reviews patient card/booklet | Nurse | Triage | Y - Refer to OPD/ART/NCD |
| **3** | | Conducts clinical assessment | Clinician | Consultation Room | Y - Order tests, diagnose |
| **4** | | Orders lab tests | Clinician | Consultation Room | |
| **5** | | Draws blood/collects sample | Lab Technician | Laboratory | |
| **6** | | Reviews lab results | Clinician | Consultation Room | Y - Confirm diagnosis |
| **7** | | Prescribes medication | Clinician | Consultation Room | |

| | | | | |
|---|---|---|---|---|
| **8** | | Dispenses medication | Pharmacist | Pharmacy |
| **9** | | Provides counseling | Counselor/Nurse | Counseling Room |
| **10** | | Schedules next appointment | Clerk/Nurse | Appointment Desk |

3. SWIMLANE 3: DATA FLOW

Purpose: To map how information is generated, captured, transmitted, and lost throughout the care process.

Instructions: Record each instance of data capture, the tools used, and whether the data was used for decision-making.

| **Step #** | **Time** | **Data Recorded** | **Data Capture Tool** | **Service Point** | **Data Used? (Y/N) & Notes** |
|---|---|---|---|---|---|
| **1** | | Demographics, vital signs | OPD Card/Register | Triage | |
| **2** | | Clinical notes, diagnosis | ART Booklet/Chronic Card | Consultation Room | Y - Reviewed past notes |
| **3** | | Lab test orders | Lab Request Form | Consultation Room | |
| **4** | | Lab test results | Lab Results Slip | Laboratory | Y - Reviewed by clinician |
| **5** | | Prescription details | Prescription Slip | Consultation Room | |
| **6** | | Medication dispensed | Pharmacy Register/EHR | Pharmacy | |
| **7** | | Next appointment date | Appointment Diary | Appointment Desk | |
| **8** | | EHR entry (OPD module) | EHR System | Data Entry Office | |
| **9** | | EHR entry (HIV module) | EHR System | Data Entry Office | |
| **10** | | Upward reporting (T5, T12) | DHIS2 | Data Entry Office | |
